# Antimicrobial resistance of bacteria isolates among patients with chronic wound infections in Tanga Regional Referral Hospital, Tanzania

**DOI:** 10.1101/2024.11.29.24318063

**Authors:** Aleena Dawer, Victor Msengi, Theresia B. Mtui, Sarah Sarakikya, Rashid Suleiman, John P. A. Lusingu

## Abstract

**Background:** Bacterial wound infections are the second leading cause of mortality globally, with approximately 50% of contaminated wounds evolving into chronic infections. In Tanzania, this challenge is exacerbated by the over prescription of antibiotics and the emergence of drug-resistant bacteria, compounded by inadequate hospital hygiene and sanitation practices. This study investigated chronic wound infections in Tanga, Tanzania, focusing on antibiotic susceptibility patterns.

**Methods:** A cross-sectional, mixed-methods study was conducted at the Tanga Regional Referral Hospital (TRRH) from July 2023 to December 2023. Pus and Culture Sensitivity tests were performed on samples from 89 chronic wound patients to identify bacterial isolates and assess antibiotic susceptibility. Data was analyzed using STATA, Excel, and Python.

**Results:** Of 89 patients, 82 (92.1%) had positive bacterial isolates in wound cultures, predominantly with *Staphylococcus aureus,* 24 (29.3%). Surgical-site infections (SSI) were the most prevalent diagnosis, followed by diabetic foot ulcers and septic wounds. Antibiotic resistance analysis revealed a marked trend in multi-drug resistance (MDR), notably against amoxicillin, while meropenem was identified as the most effective antibiotic.

**Conclusion:** The elevated rate of MDR at TRRH, particularly against commonly used antibiotics, emphasizes the need for improved antibiotic stewardship and healthcare worker education. It necessitates increased health awareness about effective wound management and the development of robust healthcare strategies to combat the escalating challenge of MDR.

## Introduction

Deaths associated with bacterial wound infections rank as the second leading cause of global mortality, according to the Global Burden of Diseases Level 3^1^. Alarmingly, approximately 50% of contaminated bacterial wounds transition into clinical infections^2^. The World Health Organization (WHO) identified hospital-acquired infections (HAI) as accounting for 25.1% of all global infections, which were attributed to surgical wounds^3^. Bacterial pathogens persist as a significant cause of morbidity and mortality worldwide^1^, posing a consistent challenge in healthcare settings. In developing nations like Tanzania, bacterial infections pose a grave threat: the incidence of wound infections is estimated to be as high as 40% as compared to the 3-11% recorded in developed countries^2^.

More specifically, *Staphylococcus aureus* (*S. aureus)* is a persistent pathogen associated with a wide range of infections, from mild skin infections to severe systemic diseases such as diabetic foot ulcers and osteomyelitis. The prevalence of *S. aureus* is rising in Tanzania, where it contributes significantly to HAI and antibiotic resistance issues^4^. The WHO’s 2017 report highlighted *S. aureus* as a priority pathogen, emphasizing the need for focused research and development of new therapeutic options due to its ability to develop resistance across multiple antibiotic classes rapidly, thereby limiting treatment options^5^.

Wound infections are highly prevalent in hospitals and often remain chronic due to improper healing. A chronic wound infection refers to a wound that fails to progress through the normal stages of healing, often remaining unhealed for more than three months. These wounds are characterized by persistent inflammation, delayed closure, and a high risk of recurring infections. Socio-economic and biological factors contribute to the prevalence of chronic wound infections, including bacterial resistance to prescribed antibiotics^6^.

Antimicrobial resistance (AMR) occurs when bacteria undergo genetic changes that allow them to withstand the effects of antibiotics that were previously effective. When patients are exposed to multiple doses of antibiotics, resistant bacteria survive and multiply, displacing the susceptible strains^7^. This resistance makes standard treatments ineffective, leading to prolonged illness, higher medical expenses, and increased mortality and morbidity. These resistant bacterial strains have spread across the globe due to the microorganism’s genetic plasticity and mobility.

This resistance is particularly high in developing countries, with underlying problems attributed to economic and societal issues, with effective antimicrobial drugs squandered by excessive use^8^. Ecological studies support the notion that higher antibiotic usage directly influences resistance^9^. This especially realms true in developing countries burdened by bacterial infectious diseases, crowded cities, poor sanitation, and the availability of antibiotics over the counter (OTC) without a prescription. These medications are often self-prescribed through unregulated supply chains^10^. Inappropriate antibiotic use, coupled with rapid disease spread through crowding and sexual contact, contributes to higher AMR rates. However, recent data remains difficult to interpret due to poor record-keeping and testing coverage in low- and middle-income countries^11^. As a result, the absence of laboratory diagnostic facilities has led to the adoption of observational diagnosis in the administration of antimicrobial medications), resulting in unnecessary and excessive treatments^12^.

In Tanzania, challenges include the easy availability of OTC antibiotics and insufficient healthcare guidance. Commonly used antibiotics in Tanzania, such as amoxicillin (34.8%), metronidazole (15.4%), tetracycline (13.3%), ciprofloxacin (6.0%), and cefalexin (4.5%), are becoming increasingly less effective due to their widespread use^13^. Moreover, antibiotic consumption is predicted to increase by 13-fold over the next twelve years in Tanzania, signaling an urgent need for action^13^.

With escalating post-operative complications and increasing antibiotic resistance in chronically infected wounds in Tanzania, understanding the bacterial spectrum and factors impairing effective treatment becomes imperative. This rationale rests significantly on the pressing need to streamline antibiotic stewardship in Tanzania. In line with WHO’s Global Antimicrobial Resistance and Use Surveillance System (GLASS) goals, which aims to foster standardized AMR data collection and analysis globally - this study contributes valuable localized data; data which will inform and shape national strategies in the fight against AMR. Given the lack of research on pre- and post-surgical chronic wound infections in the Tanga region, this data aligns with the goals of both WHO and the Tanzania National Antimicrobial Plan 2022-2028^14^.

Soaring infection rates within the surgical department is a critical health crisis that reveals gaps in healthcare stewardship within the management and prescription of wound infections. This Tanzanian research setting was vital to investigate the antimicrobial susceptibility profiles of bacterial isolates in chronic wound infections. The study not only fills existing knowledge gaps but also fosters informative practices where antimicrobial drugs are used judiciously, effectively mitigating the impact of drug-resistant infections in Tanzania and the East African region.

The research investigated the bacterial isolates and their antibiotic susceptibility patterns in patients with chronically infected wounds in the surgical ward at Tanga Regional Referral Hospital (TRRH). The study fulfilled the primary objective of identifying the bacteria responsible for chronic wound infections and their resistance patterns.

## Materials and Methods

### Study aim, design, and selection criteria

This study employed a cross-sectional approach conducted from 30/10/2023 to 03/12/2023, at TRRH. Utilizing quantitative bacterial analysis, wound swab samples were collected by the healthcare professionals in the surgical/obstetrics-gynecology, emergency, and medical-pediatric wards, employing microscopy Gram stain and culture methods to identify bacteria and their susceptibility patterns, facilitated through Pus Culture and Sensitivity tests. The cross-sectional study focused on collecting and analyzing current data, while the retrospective study reviewed past records to discern patterns and trends related to wound infections.

The selection criteria focused on all admitted patients of all ages and sexes exhibiting chronic wound infections, while excluding individuals who did not exhibit such conditions. This criterion was chosen to provide insights into the bacteria characteristics and resistance patterns prevalent in such clinical settings.

### Sample size and population

The focal point of this study was the diverse group of patients admitted to TRRH with chronic wounds. This population encapsulated both males and females, who have been afflicted with chronic wound infections that have significantly affected their quality of life.

The following were the preliminary considerations used in deriving the sample size for the quantitative study:

N*o = Sample size; Z = Confidence level; P = Estimated proportion of chronic wound infections in the population; E = Desired level of precision; Z = 1.96, P = 0.8 (80%), E = 0.05*

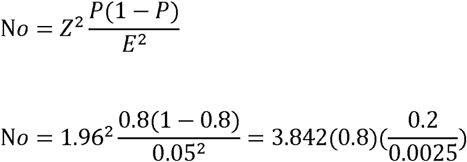

N*o* = 246 patients.

Accounting for a potential 10% loss to follow-up or refusal:

0.10 × 246 = 24.6, approximately 25 patients.

Finite population correction (Hoyle, 2015):

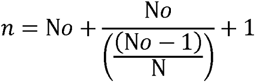

*n = Adjusted Sample size; N = Population size (100 patients);*

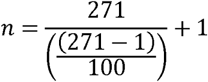

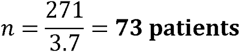

Preliminary considerations estimated the sample size to be around 73 participants. This estimation is based on trends observed in previous studies conducted in similar settings. An 80% culture positivity rate for wounds was found in the previous three months (May 2023-July 2023) at TRRH. Therefore, the actual sample size was 87 patients for quantitative analysis.

### Study setting

The Tanga Regional Referral Hospital (TRRH), famously referred to as Bombo Regional Hospital, is a significant healthcare facility located in Tanga, Tanzania. With a history dating back to the German colonial era of the 1890s, the hospital was initially established in response to the high mortality rate caused by malaria. On average, the hospital admits 28 patients per month for chronic wound infections.

### Sampling strategy

The study adopted a consecutive sampling strategy. All patients meeting the inclusion criteria during the study period were approached for sample collection until the desired sample size was achieved from 30/10/2023 to 03/12/2023.

### Ethical Statement

Ethical approval for data collection for this study was obtained from the National Health Research Ethics Review Committee (NatHREC) in Tanzania, valid from 27/10/2023 to 26/10/2024. The approval (reference: NIMR/HQ/R.8a/Vol.IX/4443) was granted for conducting research on antimicrobial susceptibility profiles in patients with chronic wound infections at Tanga Regional Referral Hospital. The study adhered to the ethical principles outlined in the Belmont Report and the Declaration of Helsinki, and progress reports were submitted to the Ministry of Health and the National Institute for Medical Research as per NatHREC guidelines. This study was conducted as part of the ‘GLOH 3950 – Global Health Practice and Research: An Experiential Learning Course’ at Georgetown University. Under this framework, local IRB approval (NatHREC) was required and obtained, while Georgetown IRB approval was not necessary for this research as per university policy. Head of Surgery at Tanga Regional Referral Hospital, Dr. Rashid Suleiman, collaborated in the study and granted permission for the work at the hospital level.

Retrospective data was collected from hospital medical records and laboratory reports covering the period 01/07/2023 to 30/10/2023. This data was accessed and analyzed between 30/10/2023 and 03/12/2023, following ethical approval. The data collection was conducted collaboratively, with Dr. Victor Msengi (Tanga Regional Referral Hospital) leading the process. Dr. Msengi anonymized all patient data before providing it to the research team, ensuring that no identifying information was accessible to the authors during or after data collection. The anonymized data included patient demographics, diagnoses, treatment history, and bacterial isolate profiles. All data handling adhered to the ethical requirements outlined by NatHREC to ensure patient confidentiality, and no attempts were made to re-identify participants.

As for prospective data collection, participants were recruited from the Tanga Regional Referral Hospital between 30/10/2023 and 03/12/2023. Eligible participants included individuals with chronic wound infections who provided consent to participate in the study. Written informed consent was obtained from all participants prior to inclusion in the study. Consent forms were available in both English and Swahili to ensure comprehension. The consent process was facilitated by Nurse Theresia Mtui, a native Swahili speaker, who distributed and explained the consent forms to participants to ensure their full understanding. No participants were recruited without written consent.

### Inclusivity in global research

Additional information regarding the ethical, cultural, and scientific considerations specific to inclusivity in global research is included in the Supporting Information.

## Data Collection Tools and Procedures

### Primary cultures

The primary goal of collecting pus samples from the wounds was to investigate the antibacterial susceptibility of the isolates present in chronic wound infections. Before collecting the pus sample, the wound sites were cleaned carefully to avoid contamination. Trained hospital staff collected pus samples using sterilized swabs, ensuring the safe and hygienic handling of samples. The collected samples were immediately transported to the laboratory in a secure transport medium to prevent any alterations in the bacterial properties. Upon receipt, swab specimens collected from the patients were systematically recorded, marking the initiation of the laboratory investigation process. This step ensures traceability and proper management of the sample data. Patient records were utilized to supplement the swab test data, filling in missing variables and providing basic clinical information such as age, gender, and clinical diagnosis. All patient information was handled in compliance with privacy and data protection regulations.

### Antibacterial Susceptibility Testing

Upon arrival to the laboratory, the samples underwent a Gram stain procedure to categorize the bacteria into Gram-positive and Gram-negative groups. Examples of Gram-positive bacteria include *Staphylococcus aureus* and *Streptococcus pneumoniae*, while *Escherichia coli and Salmonella enterica* are examples of Gram-negative bacteria. The samples were then cultured on Mueller-Hinton (MH) agar plates, setting the stage for the Kirby-Bauer disk diffusion test. The research employed antibacterial susceptibility testing using the discs recommended by the Clinical and Laboratory Standards Institute (CLSI) guidelines^15^. The study targeted both aerobic and anaerobic bacteria, given the nature of chronic wound infections, which can often be polymicrobial with both types present.

#### Preparation

6-mm filter paper disks loaded with known concentrations of antimicrobial compounds were placed on the inoculated MH agar plates.

#### Incubation

The plates were incubated at a controlled temperature of 35°C ± 2°C, facilitating bacterial growth and antimicrobial diffusion simultaneously. This process helped in establishing the zones of inhibition, where bacterial growth is curtailed by the antimicrobial agent.

#### Observation

After 24 hours, the zone of inhibition around each disk were observed. This zone size indicates susceptibility or resistance to the antimicrobial compound.

#### Recording and Interpretation

The zones of inhibition were measured, and based on the zone size, the isolates were categorized as susceptible (S), intermediate (I), or resistant (R) to the specific antimicrobial agents. This classification is done using a standardized interpretation chart based on guidelines provided by the Kirby-Bauer Disk Diffusion Susceptibility Test Protocol^16^ and TRRH’s CLSI guidelines^15^.

### Reporting

#### Data compilation

The data accumulated were meticulously recorded, documenting the susceptibility patterns of the isolates. The findings were communicated back to the clinical team, assisting them in making informed decisions regarding patient care and treatment strategies.

### Data analysis

Data analysis involved a comprehensive examination of the patterns of bacterial susceptibility and resistance. Patient records were anonymized. Statistical software was employed to analyze the data, focusing on the correlation between bacterial types, resistance patterns, and patient demographics. Data analysis was primarily conducted using STATA version 18. Microsoft Excel was employed for initial data organization and preliminary analysis. Lastly, Python played a role in the data analysis process and data visualization. This combination of STATA, Microsoft Excel, and Python provided a robust, multi-layered approach to data analysis, ensuring both the reliability and depth of the findings.

## Results

Of the 89 eligible patients, 82 had a positive wound culture (92.1%). 58.5% of these patients were male and 41.5% female. Fig 1 shows the frequency of wound infections among all age groups and sex, with a median age of 39 years and an overall range from 0-83 years. The median age for females were 35 years, with an overall range from 1-75 years. The median age for males were 39.5 years, with an overall range from 0-83 years. This suggests that wound infections were most prevalent in middle-aged patients during the study period. Most wounds were located at the lower limb, accounting for nearly 60% of all cases (n = 52). Primary diagnoses are shown in Table 1, in which 20% are attributed to surgical-site infections. However, these findings indicate a notable portion of records where wound locations were not adequately recorded.

**Fig 1.** Frequency of wound bacterial infections by age and sex at TRRH.

**Table 1.** Primary diagnoses of TRRH patients: July-November 2023 with Pus C&S Tests.

| Primary Diagnosis | Percent |
| --- | --- |
| Surgical-Site Infection | 20.9% |
| Diabetic Foot Ulcer | 18.6% |
| Otitis Media | 12.8% |
| Chronic Osteomyelitis | 11.6% |
| Septic Wound | 10.5% |
| Necrotizing Infections | 8.2% |
| Not Recorded | 5.8% |
| Abscess | 3.5% |
| Urosepsis | 3.5% |
| Pyomyositis | 2.3% |
| Pyogenic Arthritis | 2.3% |

The majority of patients had a single bacterial isolate identified in their wound (Fig 2). 80.0% of the bacterial isolates consisted of either *S. aureus, Citrobacter spp, K. pneumoniae, P. mirabilis, P. aeruginosa, E. coli* or *P. vulgaris* (Fig 2). Among the 13 distinct species identified, *S. aureus* emerged as the most prevalent, constituting nearly 29.0% of the isolates (n = 24). This prevalence underscores the significance of *S. aureus* in wound infections due to its known pathogenicity, relevance in HAI and its potential impact on treatment outcomes. *Citrobacter spp, P. mirabilis,* and *K. pneumoniae* are also notably present, each accounting for approximately 9.0% to 14.0% of the isolates, reflecting their role in clinical settings.

**Fig 2.** Distribution of bacteria isolates identified from samples at TRRH.

The Surgical Department accounted for a substantial majority of pus wound swabs (54.0%), indicative of the nature of surgical procedures, where the risk of wound infections is inherently higher due to invasive procedures and open wounds. The Emergency Medicine Department (EMD) contributed 14 swabs, making up 16.6% of the total infections. The urgency and varied nature of cases in emergency settings could explain this significant proportion. This finding highlights the areas where wound-related complications are most prevalent, guiding resource allocation and the focus of infection control initiatives.

The analysis of patient diagnoses indicated a substantial prevalence of comorbid conditions. Nearly half of the patient cohort, approximately 46.0%, were documented with multiple medical diagnoses, as opposed to a solitary primary condition. For the prevalent diagnoses of patients with infections, SSI (20.9%) and diabetic foot ulcers (18.6%) were the most common diagnoses, followed by otitis media (12.8%). Secondary and tertiary diagnoses included conditions such as diabetes mellitus, hypertension, fractures, and cellulitis. This highlights the variety of conditions leading to the need for such tests within the patient population.

The analysis of antibiotic resistance patterns in the top five bacterial isolates; *S. aureus, Citrobacter spp, K. pneumoniae, P. mirabilis, and P. aeruginosa*, (Fig 3) revealed a concerning trend in MDR. In the analysis, focus was placed on *S. aureus* due to its unique high resistance profile. Notably, amoxicillin was found to be almost universally ineffective, resisting 100.0% of all tested bacteria except *S. aureus*, exemplifying its limitation in clinical utility for this regional hospital. Similarly, cephalosporins such as ceftazidime, ceftriaxone, and cefazolin exhibited high levels of resistance in *Citrobacter, K. pneumonia,* and *P. mirabilis*, suggesting a widespread cephalosporin-resistant profile in these isolates.

**Fig 3.** Antibiogram heatmap of resistance profiles of top five bacterial isolates at TRRH.

*P. aeruginosa* had a unique pattern with complete resistance to several antibiotics, including amoxicillin, tobramycin, and augmentin, while being entirely susceptible to others like gentamycin, ciprofloxacin, and meropenem. This indicates a selective but profound AMR characteristic of this pathogen. On the other hand, meropenem emerged as the most effective antibiotic across the majority of the isolates, suggesting its potential as a first-line treatment option.

The resistance data, when viewed through the lens of the tier system^15^, underscores the necessity of strategic antibiotic selection in combating *S. aureus* infections. Tier 1 Antibiotics, considered primary for routine testing, showed varied effectiveness. For instance, cefoxitin demonstrated significant resistance (47.1%), indicating a possible prevalence of methicillin-resistant *S. aureus* (MRSA) in the isolate pool. Erythromycin exhibited an 85.7% resistance rate, suggesting limited utility in treatment. While clindamycin (38.9% resistance) might still be effective in certain clinical cases. The susceptibility of *S. aureus* to antibiotics such as meropenem, cefazolin, and tobramycin highlights effective alternative options within the local context of TRRH, especially for managing MDR strains. Additionally, the effectiveness of less commonly used antibiotics like chloramphenicol and nalidixic acid offers valuable insights for local antibiotic guidelines, particularly in resource-limited settings, reflecting unique resistance dynamics shaped by regional prescribing practices and bacterial exposure patterns in Tanga.

Tier 2 Antibiotics, used when Tier 1 antibiotics are not effective, like tetracycline showed varying resistance (38.5%). This suggests this antibiotic could be effective in MRSA cases. Other Tier 2, 3 and 4 antibiotics, reserved for high-risk MDR patients, did not appear in TRRH laboratory testing but are crucial for consideration, given their role in treating *S. aureus*.

## Discussion

These findings, underscores the complexity inherent to the patient cases encountered within the clinical setting. The implications of this data are manifold. First, it suggests that patients present with multifaceted health challenges, which necessitate a nuanced approach to treatment that extends beyond the scope of a single diagnosis. The coexistence of secondary and tertiary diagnoses alongside primary conditions, such as diabetes paired with wound infections, signals the need for integrated care pathways and a multidisciplinary approach to healthcare delivery. Furthermore, the high incidence of additional diagnoses may also reflect the interconnected nature of diseases, where one condition potentially predisposes individuals to further health complications. This pattern highlights the importance of comprehensive patient assessments to ensure that all underlying conditions are identified and addressed in the treatment plan.

Similar to previous research findings, globally, this study’s bacterial isolate profile findings have significant overlaps. Previous research studies also found that *S. aureus*^17^ was the most prevalent bacteria. This study documented *Citrobacter spp*, *Proteus mirabilis* and *K. pneumonia*e as notable contributors to wound infections as well. *P. aeruginosa* was similar in that it was one of the higher resistant isolates^18^. This study also found that meropenem was effective in bacterial isolates^19^.

This study’s results align with other studies^20,21^ done in Tanzania and Sierra Leone, showing that high Extended Spectrum β-lactamase production among Gram-negative rods contributes to widespread resistance, especially in *Citrobacter spp, K. pneumoniae,* and *P. mirabilis*. There is a clear pattern of increasing resistance to commonly used antibiotics like amoxicillin, cotrimoxazole, and cephalosporins in both studies. This trend raises concerns about the overuse and accessibility of these drugs leading to escalated bacterial resistance.

The high levels of resistance to multiple antibiotics, particularly in *S. aureus*, *Citrobacter spp, K. pneumoniae,* and *P. mirabilis*, are indicative of a burgeoning MDR crisis. These findings underscore the urgency for tailored antibiotic stewardship and the development of novel antimicrobial strategies. They also highlight the critical need for ongoing surveillance and research into antibiotic resistance patterns, which is essential for guiding effective clinical interventions and curbing the rise of MDR pathogens.

The overwhelming prevalence of swabs in the surgical department underlines the need for robust infection control measures and efficient wound management strategies in these departments. Moreover, fewer than 15.0% of swabs originated from outpatient settings. This finding supports previous research indicating a greater incidence of infections among inpatients as compared to outpatients^18^.

Compounding this concern is the prevalence of specific categories of infections observed in this study. As noted previously, SSI were found to be the most prevalent at 20.9%, closely followed by diabetic foot ulcers at 18.6%, echoing findings from previous research. For instance, Gosling^22^, identified HAI, particularly in surgical and orthopedic wards, as one of the most frequent complications, accounting for 36.7% of cases in their study titled ‘*Prevalence of hospital-acquired infections in a tertiary referral hospital in northern Tanzania*’. This correlation suggests a persistent challenge in managing SSIs across various healthcare settings. Another study’s findings^2^ on the high prevalence of polymicrobial infections in diabetic wounds, particularly involving Gram-negative rods, complement this research’s findings on the high incidence of diabetic foot ulcers.

In addition to the focus on specific categories of infections, the analysis of patient diagnoses in this study reveals a substantial prevalence of comorbid conditions. Nearly half of the patient cohort (46.0%) were documented with multiple medical diagnoses. Similar complexity has been noted in other studies, such as research on Tanzanian adults with hypertension-related diseases who used herbal treatments. In this study, 81.2% of patients presented with multiple health conditions^23^. The parallel between these findings underlines the multifaceted nature of patient health in hospital settings, where individuals often face surgical complications and other comorbid conditions, necessitating a comprehensive and integrated approach to healthcare.

The overconsumption of antibiotics and its heightened resistance also negatively impacts the economy. With increased resistance, there is an elevated cost for more expensive antibiotics, specialized equipment, longer hospital stays, and isolation procedures for patients^24^. Many patients are now resistant to first-line antimicrobials and must switch to second or third-line options, imposing a considerable strain on healthcare systems. These escalating costs might heighten the financial burden on individuals and communities, potentially leading to increased healthcare inequities^25^. This economic pressure could impede progress in improving global health outcomes, making it imperative to address antibiotic resistance with urgency and coordinated efforts across East Africa.

## Limitations

The study encountered limitations in its sample selection, specifically focusing on patients admitted to the hospital. This approach, by design, excluded the analysis of patients outside the hospital setting, potentially introducing a bias to the findings. Future studies with a more inclusive and diverse participant sample are recommended to ensure a more comprehensive understanding. For bacterial analysis, collection of swab samples was confined to patients with the financial means to afford the test costs. This financial barrier resulted in a potential overlook of a more diverse spectrum of patients from various socioeconomic backgrounds.

Another limitation was the unavailability of a full spectrum of antibiotics for testing in the laboratory due to resource constraints. To maintain laboratory protocol, testing was guided by Clinical and Laboratory Standards Institute (CLSI) guidelines, which recommend specific antibiotics for different bacterial isolates, leading to some variability in the antibiotics tested across samples. Future studies should secure sufficient funding to ensure that testing costs are not a hindrance. By doing so, future research can provide a more inclusive dataset, capturing a wider range of patient bacterial profiles and thereby enriching the understanding of the subject matter. Moreover, while swabbing is a commonly used, non-invasive, and cost-effective method for specimen collection, it does have inherent drawbacks. Inadequate specimen collection has the potential to skew results. However, in this study, this potential limitation was reduced by thoroughly cleansing the wound before swab collection, thereby reducing the likelihood of such contamination.

Lastly, the small sample size limited the capacity to perform advanced statistical analyses that could have enhanced the study’s robustness. Regarding generalizability, the findings provide localized data specific to TRRH rather than reflecting broader population trends, which may limit external validity. Additionally, due to the study’s limited resources, the data does not differentiate between community-associated and healthcare-associated infections. Limited information from healthcare providers about patients’ prior antibiotic use, particularly OTC purchases before seeking hospital care, further constrained additional extrapolative analysis. This limitation complicates understanding the timing of antibiotic use, as many patients potentially self-medicate prior to seeking professional care. Future studies might address this by including community data and capturing antibiotic purchase histories to improve the context of AMR patterns.

## Recommendations

The high prevalence of inadequate care is deeply intertwined with broader social determinants of health, including poverty, education, and social stratification. These factors collectively play a pivotal role in shaping healthcare accessibility and quality. In light of these findings, the study proposes two main recommendations. These recommendations are designed to be practical and actionable, addressing the urgent needs identified in the research. Their implementation possesses the potential to significantly improve patient outcomes, alleviate the burden of AMR and enhance overall quality of healthcare delivery in the region.

### Strengthening surveillance on antibiotic resistance

Enhancing surveillance systems and research efforts is crucial for understanding and monitoring the trends and patterns of antibiotic resistance. This would aid in developing targeted strategies to mitigate the spread of MDR pathogens. This recommendation involves expanding the capability of both hospital-based and community-based surveillance networks to accurately track trends and patterns of antibiotic resistance. The logistics of this enhancement would include equipping hospitals and community health centers with the necessary technology and training staff to collect, analyze, and report data on antibiotic use and resistance patterns. This data would provide a clearer picture of the resistance landscape, enabling more targeted and effective interventions.

Another key element in this strategy is fostering collaboration between academic institutions, healthcare facilities, and government health agencies. This collaborative effort would focus on conducting in-depth research to understand the causes, mechanisms, and spread of MDR. By pooling resources and expertise, these institutions can undertake more comprehensive and sophisticated research projects. Such collaboration could involve sharing samples, data, and research findings, as well as jointly applying for research grants and funding. This cooperative approach would not only enhance the quality of the research, but also expedite the process of gaining critical insights into antibiotic resistance.

Establishing a system for regular data sharing and reporting among healthcare facilities and public health agencies is crucial. This system would facilitate a coordinated response to MDR challenges by providing all stakeholders with timely and accurate information. This could be achieved through the creation of a centralized database accessible to relevant parties, where data on antibiotic use and resistance can be regularly updated and analyzed. Regular meetings and conferences could also be organized to discuss findings, share best practices, and coordinate efforts.

Based on the surveillance data and research findings, targeted public health interventions can be implemented. These interventions might include infection control measures in hospitals and public awareness campaigns about the importance of responsible antibiotic use. Tailoring these interventions to specific communities based on the data collected ensures that they are as effective as possible. Moreover, these interventions would need to be dynamic, adapting to the evolving nature of antibiotic resistance.

As mentioned prior, the GLASS-AMR WHO action plan calls for nations to develop standardized AMR data collection and analysis based on regional data. The findings from the Tanga region study directly support this goal, offering insights that are region-specific and therefore more relevant and actionable for local health authorities. The Tanzania National Antimicrobial Plan 2022-2028 also emphasizes the need for localized research and data to inform its strategies against AMR. This alignment significantly enhances the potential for securing funding, as the study’s objectives align with those of major global and national health initiatives. Securing funding for the proposed enhancements in surveillance systems, research collaborations, and public health interventions becomes more feasible when the research is aligned with these larger global and national initiatives.

### Development of community-based health programs

Transitioning from the broader context of AMR to the more focused realm of wound infection management, the development of community-based health programs emerges as a crucial recommendation. These programs, aimed at early detection and management of wound infections, should be made accessible and affordable to all patients, regardless of their socioeconomic status. This accessibility can be facilitated through subsidies from government or non-governmental organizations. The program would implement mobile clinics in rural and underserved areas within Tanzania. These clinics will provide essential services such as basic wound care, education on wound management, and early detection of potential complications. Their mobility ensures that healthcare reaches those who are geographically isolated or lack access to traditional healthcare facilities. Complementing mobile clinics, regular health workshops in local communities are crucial. These workshops will serve as platforms to educate residents about the importance of proper wound care and the role of nutrition in the healing process. By raising awareness, these workshops can empower individuals to take proactive steps in managing their health and seeking timely medical intervention.

An integral part of this recommendation is the training and deployment of community HCWs. This aspect is particularly crucial in light of the disparities in HCW knowledge observed in the study. These HCWs, equipped with the necessary skills and knowledge, can conduct home visits to patients who are immobile or lack access to healthcare facilities. Their role extends beyond providing basic wound care; they are also educators and advisors to patients and their families. They can impart crucial knowledge about wound management and prevention strategies, ensuring that even the most vulnerable sections of the community have access to healthcare information. These community health workers would act as a vital link between the patients and the healthcare system. They can identify cases that require more advanced medical attention and facilitate referrals to hospitals, ensuring that patients receive comprehensive care.

## Conclusions

The study emphasized the urgent challenge of AMR, especially among pathogens such as *S. aureus, Citrobacter spp, K. pneumoniae,* and *P. mirabilis.* Addressing this issue requires immediate action in antibiotic stewardship and the development of novel treatment strategies. The study revealed significant MDR and high infection rates particularly in the surgical ward. However, it also identifies opportunities for targeted interventions, emphasizing the need for enhanced public health and healthcare worker education, improved healthcare infrastructure, and integrated approaches that respect cultural contexts.

This study’s unique contribution lies in its exploration of AMR at a Tanzanian regional referral hospital, highlighting critical areas for intervention and improvement in healthcare strategies. These insights are especially vital in settings where resource constraints significantly influence health management. The recommendations proposed herein aim to improve patient outcomes, reduce the burden of AMR, and enhance the overall quality of healthcare delivery in the local context. In doing so, they contribute to the global effort in managing and preventing chronic wound infections.

## Declarations

### Ethics approval and consent to participate

The proposed study is grounded in the ethical principles of beneficence, non-maleficence, and respect for autonomy. The anticipated benefits of the study, which include enhancing knowledge about chronic wound infections and potentially developing more effective treatment strategies, justify the ethical conduct of the study. The study protocol was endorsed through rigorous scientific ethical review by the National Institute of Medical Research (NIMR) (ethical approval NIMR/HQ/R.8a/Vol. IX/4443 granted on 27/10/2023). All participants signed a written informed consent prior to inclusion. All methods were performed in accordance with the relevant guidelines and regulations.

### Consent for publication

I hereby provide consent for the publication of the manuscript detailed above, including any accompanying images or data contained within the manuscript.

### Availability of data and materials

The datasets used and/or analysed during the current study are available in the Dryad repository. The data can be accessed at the following

DOI: http://datadryad.org/stash/share/o8XJQKJH7tVM6SnBA5wY0K9-nV9S5VZc0MCY52h24V4.

## Funding

This research was funded by Georgetown University’s Global Health Department.

### Authors’ contributions

Principal investigator: AD; Study design: AD, JPAL, VM; Data collection: TM, VM, AD; Laboratory procedures: SS, TM; Data analysis: AD, JPAL; Approved hospital use: RS; wrote draft of manuscript: AD; and reviewed and approved the final draft of the manuscripts: JPAL, AD, VM, SS, TM, RS.

## Data Availability

The dataset underlying this study has been fully anonymized to protect patient confidentiality and has been made available in the Dryad repository. It can be accessed at https://doi.org/10.5061/dryad.z08kprrq3. The dataset includes variables related to patient demographics, diagnoses, antimicrobial resistance patterns, and hospitalization durations. All personally identifiable information has been removed to ensure compliance with ethical guidelines.

https://datadryad.org/stash/share/o8XJQKJH7tVM6SnBA5wY0K9-nV9S5VZc0MCY52h24V4

## Acknowledgments

We thank all patients, laboratory scientists & medical microbiologists and medical staff at Tanga Regional Referral Hospital, Tanzania and the Institute of Medical Research for their assistance and continuous support.

## References

[1] World Health Organization. Report signals increasing resistance to antibiotics in bacterial infections in humans and need for better data. WHO 2022b. https://www.who.int/news/item/09

[2] Kassam, N.A., Damian, D.J., Kajeguka, D. et al. Spectrum and antibiogram of bacteria isolated from patients presenting with infected wounds in a Tertiary Hospital, northern Tanzania. BMC Res Notes 2017; 10, 757. 10.1186/s13104-017-3092-9

[3] Tikhomirov, E. WHO programme for the control of hospital infections. Chemioterapia 1987; 6(3), 148–151.

[4] Mzee T, Kazimoto T, Madata J, et al. Prevalence, antimicrobial susceptibility and genotypic characteristics of *Staphylococcus aureus* in Tanzania: a systematic review. Bull Natl Res Cent. 2021;45:162. doi:10.1186/s42269-021-00612-z

[5] World Health Organization. Global priority list of antibiotic-resistant bacteria to guide research, discovery, and development of new antibiotics. Geneva: WHO; 2017.

[6] Ventola C. L. The antibiotic resistance crisis: part 1: causes and threats. P & T 2015; 40(4), 277–283.

[7] Habboush Y, Guzman N. Antibiotic resistance. [Updated 2023 Jun 20]. In: StatPearls. Treasure Island (FL): StatPearls Publishing; 2024 Jan. https://www.ncbi.nlm.nih.gov/books/NBK513277/

[8] Kunin C. M. Resistance to antimicrobial drugs--a worldwide calamity. Ann. Intern. Med. 1993; 118(7), 557–561. 10.7326/0003-4819-118-7-199304010-00011

[9] Goossens H. Antibiotic consumption and link to resistance. CMI: ESCMID 2009; 15 Suppl 3, 12–15. 10.1111/j.1469-0691.2009.02725.x

[10] Ayukekbong, J.A., Ntemgwa, M. & Atabe, A.N. The threat of antimicrobial resistance in developing countries: causes and control strategies. Antimicrob Resist Infect Control 2017; 6, 47. 10.1186/s13756-017-0208-x

[11] World Health Organization. Global antimicrobial resistance and use surveillance system (GLASS) report: 2022a. https://www.who.int/publications/i/item/9789240062702

[12] Wall S. Prevention of antibiotic resistance - an epidemiological scoping review to identify research categories and knowledge gaps. GHA 2019; 12(1), 1756191. 10.1080/16549716.2020.1756191

[13] Sangeda, R. Z., Saburi, H. A., Masatu, F. C., et. al. National antibiotics utilization trends for human use in Tanzania from 2010 to 2016 inferred from Tanzania medicines and medical devices authority importation data. Antibiotics 2021; (Basel, Switzerland), 10(10), 1249. 10.3390/antibiotics10101249

[14] Ministry of Health, United Republic of Tanzania. The United Republic of Tanzania National Action Plan on Antimicrobial Resistance 2023-2028. World Health Organization; 2023. Available from: https://cdn.who.int/media/docs/default-source/antimicrobial-resistance/amr-spc-npm/nap-library/the-united-republic-of-tanzania---national-action-plan-2023--2028.pdf?

[15] CLSI Clinical & Laboratory Standards Institute. CLSI M100 performance standards for antimicrobial susceptibility testing. 33rd ed. CLSI 2022. https://clsi.org/standards/products/microbiology/documents/m100/

[16] Hudzicki, J. Kirby-Bauer Disk Diffusion Susceptibility Test Protocol. ASM 2009.

[17] Mnyambwa NP, Mahende C, Wilfred A, Sandi E, Mgina N, Lubinza C, et al. Antibiotic susceptibility patterns of bacterial isolates from routine clinical specimens from referral hospitals in Tanzania: a prospective hospital-based observational study. Infect Drug Resist. 2021;14:869–878. doi:10.2147/IDR.S294575. PMID: 33688222; PMCID: PMC7937390.

[18] Dekker, D., Pankok, F., Thye, T., Taudien, S., Oppong, K., Akenten, et. al. Clonal clusters, molecular resistance mechanisms and virulence factors of Gram-Negative bacteria isolated from chronic wounds in Ghana. Antibiotics 2021; (Basel, Switzerland), 10(3), 339. 10.3390/antibiotics10030339

[19] Agyepong, N., Govinden, U., Owusu-Ofori, A., & Essack, S. Y. Multidrug-resistant gram-negative bacterial infections in a teaching hospital in Ghana. Antimicrob Resist Infect Control 2018; 7, 37. 10.1186/s13756-018-0324-2

[20] Mloka, D., Sangeda, R. Z., Mwambete, K. D., & Kamuhabwa, A. R. Magnitude of Extended-Spectrum Beta-Lactamase-Producing Gram-Negative and Beta-Lactamase-Producing Gram-Positive pathogens isolated from patients in Dar es Salaam, Tanzania: A cross-sectional study. Cureus 2022; 14(4), e24451. 10.7759/cureus.24451

[21] Schaumburg, F., Vas Nunes, J., Mönnink, G., Falama, A. M., Bangura, et. al. Chronic wounds in Sierra Leone: pathogen spectrum and antimicrobial susceptibility. Infection 2022; 50(4), 907–914. 10.1007/s15010-022-01762-6

[22] Gosling, R., Mbatia, R., Savage, A., Mulligan, J. A., & Reyburn, H. Prevalence of hospital-acquired infections in a tertiary referral hospital in northern Tanzania. Ann Trop Med Parasitol 2003; 97(1), 69–73. 10.1179/000349803125002724

[23] Liwa, A., Roediger, R., Jaka, H., et. al. Herbal and alternative medicine use in Tanzanian adults admitted with hypertension-related diseases: A mixed-methods study. Int. J. Hypertens 2017; Article 5692572. 10.1155/2017/5692572

[24] Prestinaci, F., Pezzotti, P., & Pantosti, A. Antimicrobial resistance: a global multifaceted phenomenon. PGH 2015; 109(7), 309–318. 10.1179/2047773215Y.0000000030

[25] Janssen, J., Afari-Asiedu, S., Monnier, A. et al. Exploring the economic impact of inappropriate antibiotic use: the case of upper respiratory tract infections in Ghana. Antimicrob Resist Infect Control 2022; 11, 53. 10.1186/s13756-022-01096-w

